# Plasma from recovered COVID19 subjects inhibits spike protein binding to ACE2 in a microsphere-based inhibition assay

**DOI:** 10.1101/2020.06.09.20127050

**Authors:** Edward P. Gniffke, Whitney E. Harrington, Nicholas Dambrauskas, Yonghou Jiang, Olesya Trakhimets, Vladimir Vigdorovich, Lisa Frenkel, D. Noah Sather, Stephen E.P. Smith

## Abstract

High throughput serological tests that can establish the presence and functional activity of anti-SARS-COV2 antibodies are urgently needed. Here we present microsphere-based Flow Cytometry assays that quantify both anti-spike IgGs in plasma, and the ability of plasma to inhibit the binding of spike protein to angiotensin converting enzyme 2 (ACE2). First, we detected anti-spike-trimer IgGs in 22/24 and anti-spike-receptor-binding-domain (RBD) IgGs in 21/24 COVID+ subjects at a median of 36 (range 14-73) days following documented SARS-CoV-2 RNA (+) secretions. Next, we find that plasma from all 22/24 subjects with anti-trimer IgGs inhibited ACE2-trimer binding to a greater degree than controls, and that the degree of inhibition correlated with anti-trimer IgG levels. Depletion of trimer-reactive Igs from plasma reduced ACE2-trimer inhibitory capacity to a greater degree than depletion of RBD-reactive Igs, suggesting that inhibitory antibodies act by binding both within and outside of the RBD. Amongst the 24 subjects, presence of fever was associated with higher levels of anti-trimer IgG and inhibition of binding to human ACE2. This inhibition assay may be broadly useful to quantify the functional antibody response of recovered COVID19 patients or vaccine recipients in a cell-free assay system.

## Introduction

In late 2019 a novel SARS-like coronavirus (SARS-CoV-2) emerged in Wuhan, China^1^. It rapidly spread across the globe, and the disease it causes, COVID19, was declared a pandemic by the WHO in March 2020^2^. The cryo-EM structure of the spike glycoprotein, a viral surface protein that mediates cell entry of corona viruses, was solved by two groups^3,4^, revealing a trimeric structure with either none or one of three receptor binding domains (RBDs) in the “up” state, capable of binding to its target. Similar to the 2003 SARS-CoV-1 virus, angiotensin converting enzyme 2 (ACE2) serves as the receptor necessary and sufficient for infection of the target cell^3–7^.

Nearly all patients who recover from SARS-CoV-2 infection produce IgM and IgG antibodies against the spike protein^7–9^, and a large number of serologic tests have been produced and marketed. However, it is unclear how effective the detected antibodies are at neutralizing viral activity. The percentage of people who show functional responses using cell-or protein-based neutralizing assays varies depending upon the analytical strategy used; for example a study measuring in vitro inhibition of ACE2-RBD binding in an ELISA-style assay showed only 3 of 26 (11.5%) recovered patients strongly inhibited binding^7^, while a larger study using pseudotyped lentivirus showed significant inhibition of infectivity in 165 of 175 (94.3%) recovered patients^9^. Indeed, human monoclonal antibodies that neutralize SARS-CoV-2 infectivity have been show to bind epitopes both within^7^ and outside of the RBD^10^, so it is unclear if RBD-based measurements capture the full repertoire of inhibition of viral infectivity. While pseudoviruses that use the native spike protein to infect cells appear to at identify antibodies that neutralize viral entry^9^, these assays are technically demanding, require specialized biosafety facilities, and may be difficult to scale up for population-level testing.

Here, we describe a novel, overnight, cell-free assay to quantify a plasma sample’s ability to inhibit the binding of ACE2 to the recombinant trimeric spike protein^3^. The assay is based on immunoprecipitation detected by flow cytometry (IP-FCM) technology, a highly sensitive and reagent efficient method for detecting protein-protein interactions using minimal amounts of biomaterial^11,12^. We find that while only a minority of persons who have recovered from symptoms of COVID-19 produce antibodies that inhibit the RBD domain binding to ACE2 in vitro, almost all (22/24) produce antibodies that potently inhibit the full trimer binding to ACE2. Our results provide a new, relatively rapid and high-throughput method to quantify circulating levels of functional anti-SARS-CoV-2 antibodies, and suggest that the entire spike protein as opposed to the RBD should be used when characterizing SARS-CoV-2 immunity.

## Results

### IgG Detection

We designed and optimized IP-FCM assays to detect the presence of anti-SARS-CoV-2 spike antibodies in human serological samples by covalently coupling either the recombinant RBD fragment^13^ or a spike trimer construct^3^ to carboxy-modified polystyrene latex (CML, data shown in supplemental figures) or to Luminex MagPlex microspheres. Coupled beads were then incubated with increasing dilutions of plasma specimens, and then with anti-human IgG antibodies conjugated to phycoerythrin (PE) (Figure 1A). Plasma specimens were obtained from participants with SARS-CoV-2 RNA (+) nasopharyngeal secretions detected by rt-PCR at ≥14+ days post-symptom onset. These participants include COVID19+ subjects, confirmed by RTPCR, enrolled into the Seattle Children’s SARS2 Recovered Cohort on/by 6/1/2020. Additionally, de-identified banked plasmas from healthy individuals collected before 2019 were used as negative controls. A plasma-dilution experiment using specimens from three SARS-CoV-2 RNA (+) and two pre-2019 samples detected high levels of anti-human IgG in the positive samples, and non-specific background reactivity in the controls that decreased to near 0 at higher dilutions (Figure S1A,B). Note that when using IP-FCM, lower median fluorescent intensity (MFI) values at high concentrations of analyte can occur, resulting the U-shaped curve shown in Figure S1A. We chose 1:1000 as an optimal plasma dilution, and separately identified 2 hours at room temperature (22-24°C) as the incubation conditions that resulted in the best signal-to-noise ratio (data not shown).

**Figure 1:**
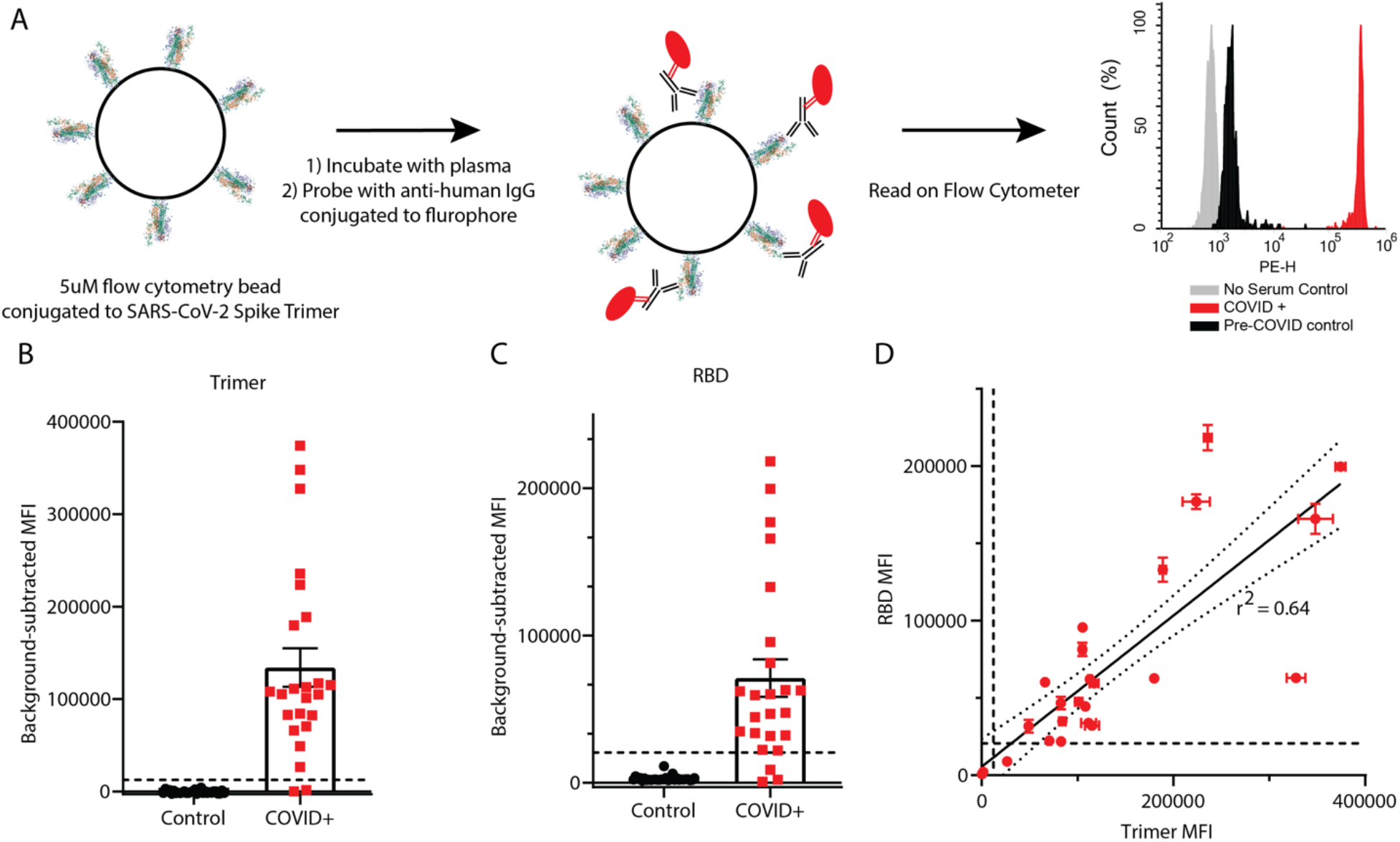
Detection of anti-SARS-CoV-2 IgG using RBD and trimer constructs. (A) Graphical representation of the methods. (B, C) IgG levels in plasma from 24 COVID+ and 30 pre-COVID-19 subjects were measured using trimer-conjugated (B) or RBD-conjugated (C) microspheres. Dashed lines indicate the cut-off for positive sample designation, calculated as (Maximum control value + 5 standard deviations, at MFI = 12,432 for trimer, 27119 for RBD. E) The median fluorescent intensity of IgG measured on the trimer and RBD assays was significantly correlated. Dashed lines indicate cut-offs for positive designation, and the solid line shows a linear regression with 95% confidence intervals indicated with dotted lines, r^2^ = 0.64, slope is significantly non-zero (F_1,46_=82.4, p<0.0001).

Using MFI from the 30 control specimens, SARS-CoV-2 antibodies were stringently defined as “positive” at an MFI greater than the highest control value plus 5 standard deviations of the control values (dashed lines in Figure 1B,C). Using this definition, 22/24 (91.6%) COVID+ samples were classified as ‘seropositive’ for trimer reactivity, and 21/24 (87.5%) for RBD. There was a moderate correlation between RBD and trimer immunoreactivity (Figure 1D, r^2^ = 0.64). Experiments performed using CML microspheres during assay development using a subset of the total plasma samples showed similar results (Figure S1), and there was correlation between trimer (r^2^ = 0.85) and RBD (r^2^ = 0.75) values for the 12 samples that were measured using both microsphere types (Figure S1F, G).

### Inhibition of ACE2 binding

To determine if the presence of plasma IgG might correlate with inhibition of the virus binding to its receptor, we designed an IP-FCM assay to quantify the binding of ACE2 and SARS-CoV-2 spike protein. We coupled human ACE2 protein to CML (Figure S2) or MagPlex (Figure 2) microspheres and incubated with biotinylated RBD or trimer, followed by incubation with Streptavidin-PE (Figure 2A). Both trimer and RBD demonstrated strong binding to ACE2 (Figure 2B,C), and the EC_50_ was used to determine the dilution of spike proteins added to the binding inhibition assay. As a specificity control, unlabeled human ACE2 inhibited binding of both RBD and trimer by >90% (Figure 2D,E). After optimizing the plasma dilution (Figure S2C,D,), we evaluated the same 24 COVID19+ and 30 pre-COVID patient plasma samples for their ability to inhibit the ACE2-Spike interaction. Analyzed as a group, the median fluorescent intensity (MFI) of control plasma was not significantly different than no-plasma controls in the trimer or RBD assays (Figure 2F or H, respectively), while COVID+ samples significantly inhibited binding (trimer ANOVA F_(2,57)_=52.4, p<0.0001, COVID+ vs. no plasma and pre-COVID p<0.0001 by Tukey post-hoc test; RBD ANOVA F_(2,57)_=5.538, p=0.006, COVID+ vs. pre-COVID p = 0.004 by Tukey post-hoc test). Similarly, when data were expressed as percent inhibition, COVID+ samples produced greater inhibition than pre-COVID for both trimer and RBD (p<0.0001 and p=0.0024, respectively, by 2-tailed t-test, Figure 2 G,I).

**Figure 2:**
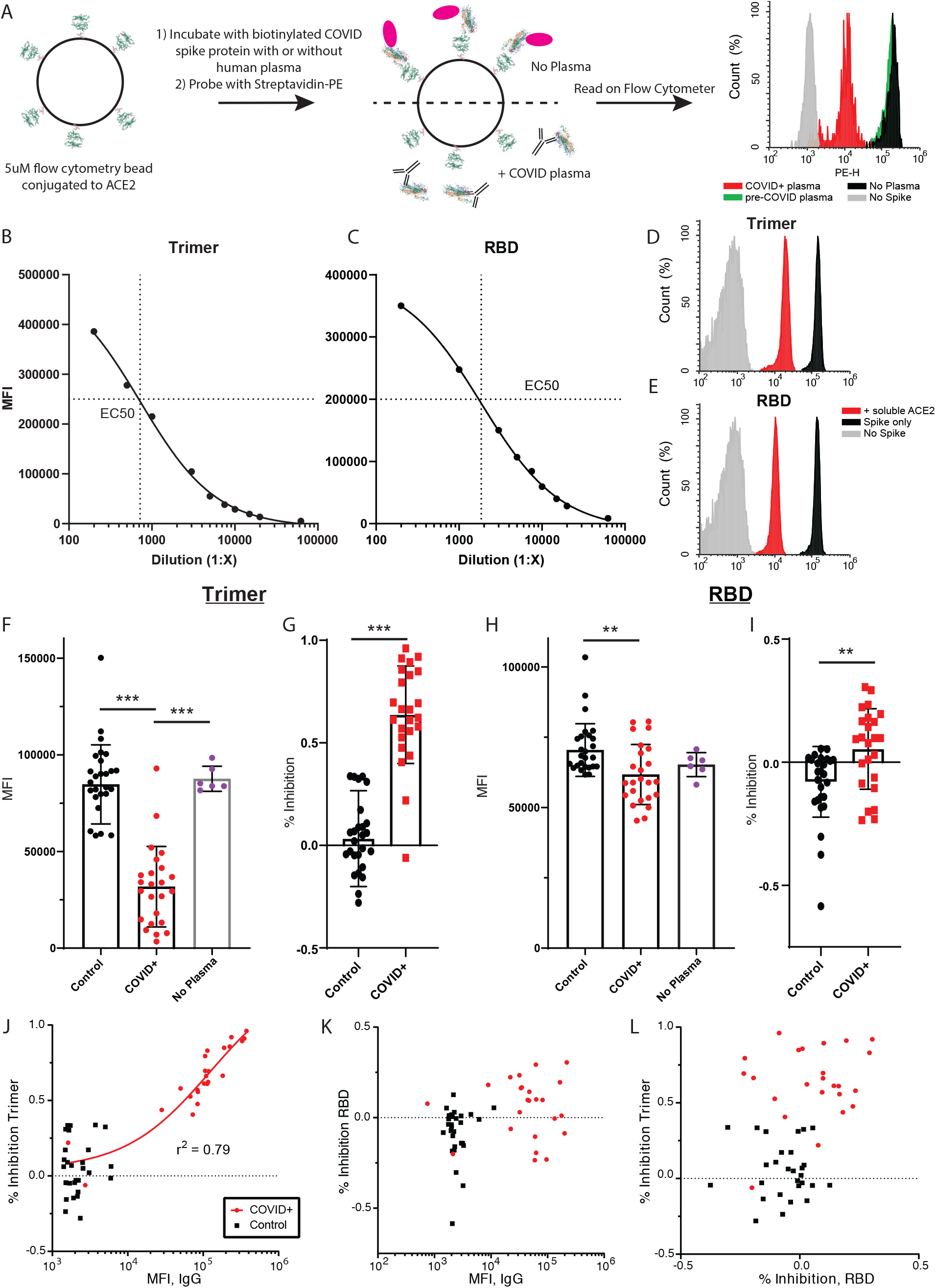
Inhibition of SARS-CoV-2 spike protein and ACE2 binding by recovered patient plasma. (A) Graphical representation of the methods. (B, C) The EC50 of the Ace2-Trimer (B) or Ace2-RBD (C) binding reaction was determined by serial dilution of the trimer or RBD fragment. (D,E) Soluble, unlabeled ACE2 was added to the trimer (D) or RBD (E) binding assay (red histograms), which resulted in a >90% reduction in MFI compared to binding assays without soluble Ace2 (black histograms). RBD or trimer was omitted to establish background MFI (grey histograms). (F) Compared to a no-plasma control (purple) or pre-COVID control plasma (black), COVID+ plasma significantly inhibited trimer-ACE2 binding *** p<0.0001. (G) Same data as in F, expressed as % inhibition vs the no-plasma control mean. *** P<0.0001. (H) Compared to pre-COVID plasma (black) COVID+ plasma significantly inhibited RBD-Ace2 binding (but not compared to a no-plasma control (purple)). ** p = 0.004. (I) Same data as in C expressed as % inhibition vs the no-plasma control mean. ** p = 0.0024. J) Trimer IgG assay MFI plotted vs. % inhibition on the Trimer-Ace2 inhibition assay for all samples. A regression line for all COVID+ samples is shown. K) RBD IgG assay MFI plotted vs. % inhibition on the RBD-Ace2 inhibition assay for all samples. L) % inhibition on the trimer-Ace2 inhibition assay plotted vs. % inhibition on the RBD-Ace2 inhibition assay.

However, when data were examined by individual, important differences emerge between the trimer and RBD-based assays. For the trimer inhibition assay, all 22 plasma samples that tested positive for anti-trimer IgG (as defined above) showed greater inhibition than any of the negative control samples (Figure 2J). Conversely, in the RBD assay, only 9/24 samples showed greater inhibition than the highest negative control, despite 21/24 samples being sero-positive (Figure 2K). Moreover, in the trimer assay there was a strong correlation between the amount of anti-trimer IgG and the percent inhibition among COVID+ (r^2^ = 0.79) but not negative control (r^2^ = 0.04) samples (Figure 2J). There was no such correlation in the RBD assay (COVID+ r^2^ = 0.047, negative control r^2^ = 0.034, Figure 2K). There was also no correlation between the percent inhibition of plasma samples in the RBD and trimer assays (Figure 2L, COVID+ r^2^ = 0.036, pre-COVID r^2^ = 0.011). For the two Trimer-negative samples, follow-up testing using the commercial Cellex platform as well as laboratory-developed ELISAs using spike and nucleoprotein (n=3 different labs) failed to detected anti-SARS2 antibodies.

Experiments performed using CML microspheres during assay development using a subset of the total plasma showed similar results (Figure S2). There was good correlation between trimer values for the 12 samples that were measured using both microsphere types (r^2^ = 0.85, Figure S2K), and MagPlex microspheres were more effective at distributing the population across the dynamic range of the assay. For the RBD assay, there was poor correlation between values measured using both microsphere types (r^2^ = 0.15, Figure S2L), but samples that were more effective at inhibition appeared to show a trend toward a linear relationship. Overall our data suggest that the majority of participants produces anti-trimer antibodies that efficiently disrupt ACE2-trimer binding; but while most participants also produce anti-RBD antibodies, less than half also disrupt ACE2-RBD binding.

### Antibody depletion assay

We next depleted the plasma of one negative control and three COVID+ samples by incubating plasmas with large numbers of trimer- or RBD-conjugated CML beads for three consecutive overnight incubations until the levels of IgG detected in the plasma plateaued (Figure 3A, B). We then ran the trimer-ACE2 inhibition assay on the depleted plasma to determine if inhibition of binding was caused by antibodies binding within or outside of the RBD. Prior to depletion, the COVID+ samples showed varying levels of ACE2-trimer binding inhibition (41-75%). Inhibition was reduced by partially (although non-significantly) reduced by RBD depletion, and was reduced to a greater degree by trimer depletion (Figure 3C, ANOVA F_(2,6)_=8.51, p=0.018, Not depleted vs. RBD depleted p = 0.11, Not depleted vs. trimer depleted p = 0.015, RBD depleted vs. trimer depleted = 0.30 by Tukey post-hoc test). Thus, even though RBD depletion was more efficient than trimer depletion in terms of reduction in detectable IgG (RBD: X%, trimer Y%, Fig 3A,B), trimer depletion was more at removing the neutralizing capacity of the plasma. These data suggest that spike trimer neutralizing antibodies bind both within and outside of the RBD.

**Figure 3:**
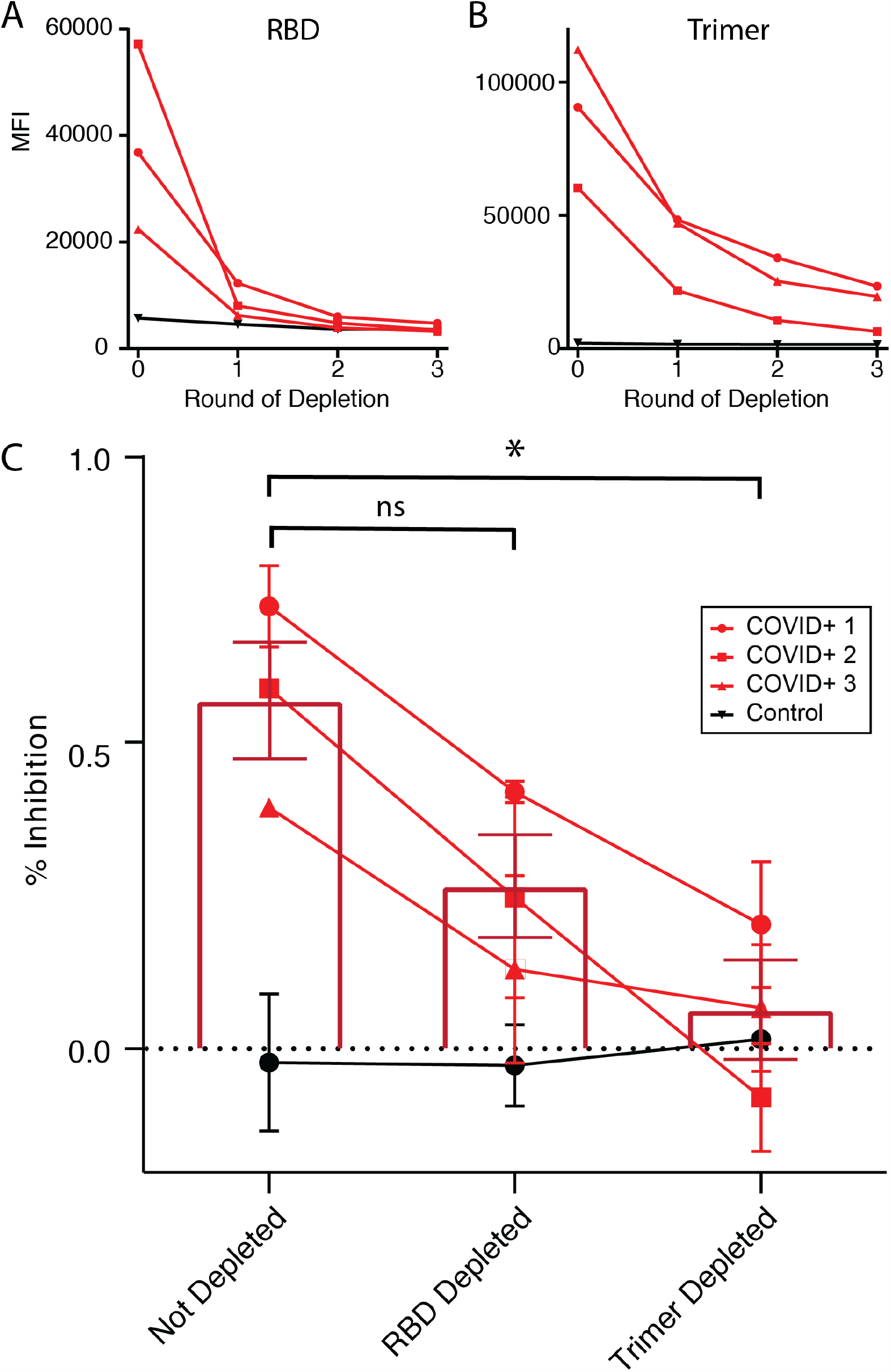
Depletion of trimer-binding IGs prevents inhibition of Ace2-trimer binding. A,B) Plasma samples from three COVID+ and one control individual were depleted of RBD-binding Igs (A) or trimer-binding Igs (B) by incubation with three rounds of RBD- or trimer-conjugated CML beads. After each round, a sample was removed and assayed for remaining anti-RBD or anti-trimer IgG. C) COVID+, not-depleted plasma inhibited trimer-ACE2 binding. After RBD-depletion, plasma was less efficient at inhibiting trimer-ACE2 binding, although the difference was not significant by ANOVA (see text for statistics). After trimer-depletion, the ability of plasma to inhibit ACE2-trimer binding was significantly reduced, with the mean overlapping with control levels. ANOVA F_(2,6)_=8.51, p=0.018, Not depleted vs. RBD depleted p = 0.11, ns; Not depleted vs. trimer depleted * p = 0.015; RBD depleted vs. trimer depleted = 0.30 by Tukey post-hoc test.

### Correlations between COVID+ subject characteristics and serology results

Participants (n=24) were between 28 and 63 years of age (median 42 years of age) and were 71% (17/24) female. The day of sample collection ranged from 14 to 73 days from symptom onset, with a median of 36 days. All SARS-CoV-2 infections were symptomatic, ranging from very mild to moderate illness; no subjects required hospitalization or supplemental oxygen. 12/24 subjects experienced fever and 16/24 experienced cough. Age was associated with a trend toward increased anti-trimer IgG and inhibition (per year MFI difference: 3,816, p=0.09; inhibition difference: 1%, p=0.07), although this may have been driven by the two subjects with negative IgG, who were both relatively young (28, 41). Sex, days since symptom onset, and cough were not associated with anti-trimer IgG or inhibition. In contrast, fever was strongly associated with both anti-trimer IgG and inhibition in both unadjusted (MFI difference: 103,982, p=0.007; inhibition difference: 20%, p=0.04) and adjusted models that accounted for subject age, sex, and days from symptom onset (adjusted MFI difference: 103,530, p=0.01; adjusted inhibition difference: 20%, p=0.03). Both IgG-negative subjects had very mild disease without fever.

## Discussion

The acquisition of antiviral immunity to SARS-CoV-2 will be a critical prerequisite for a return to normalcy from the current pandemic. As of this writing, it is still unclear if infection or vaccination will lead to robust, long-lasting immunity^14^. While some early data suggested ineffective immune responses, such as low levels of ACE2-RBD inhibition by convalescent serum in an ELISA-style binding assay^7^ or the detection of viral RNA in recovered patients^15^, more recent data suggest effective anti-viral antibody activity in the majority of patients. For example, serum from 94% of recovered patients effectively inhibited pseudotyped lentiviral infection of susceptible cells^9^, similar to our rates; four Rhesus Macaques experimentally infected with SARS-CoV-2 were resistant to re-infection^16^; and a recent report suggests that the viral RNA detected in recovered patients is not associated with an active infection or the ability to transmit COVID19^17^. Our data, demonstrating in vitro neutralization of ACE2-trimer binding in 92% of recovered patients, adds to this growing literature that collectively suggest that infection with SARS-CoV-2 results robust immunity, at least in the short-term.

While we identified robust trimer-ACE2 inhibition in IgG-positive post-convalescent plasma, only weak inhibition of RDB-ACE2 binding was detectable, and only in the minority of samples. Moreover, there was no correlation between anti-RBD levels and inhibition of ACE2-RBD binding, suggesting that some people may mount an antibody response to the RBD that fails to inhibit binding of the RBD construct to recombinant, bead-bound ACE2. However, this failure may be related to the relative affinity of the RBD construct to ACE2 vs. the affinity of plasma antibodies to the RBD, and thus not be relevant to in vivo immunity. Indeed, depletion of anti-RBD antibodies reduced the trimer-inhibitory activity of plasma, suggesting that at least some trimer-inhibitory antibodies bind within the RDB. This is consistent with published studies of monoclonal antibodies, which have been found to bind both within^7^ and outside of^10^ the RBD. Regardless of mechanism, we report that for practical purposes, the trimer construct provides a reliable, quantitative readout of immunity while the RBD construct is less useful.

Our cohort was comprised of relatively young, healthy subjects, none of whom were hospitalized or required supplemental oxygen. Amongst these individuals, the levels of anti-trimer IgG and inhibition were higher in those that experienced fever versus no fever. Findings regarding correlation between antibody levels and clinical severity have been mixed^18,19^, with one study describing higher antibody levels in severe versus moderate COVID19 cases, all of whom were hospitalized^8^. To the best of our knowledge, this is the first description of a correlation between illness severity and antibody levels in a community cohort. Our findings may suggest that systemic illness or an inflammatory response lead to higher antibody titer; alternatively, both the presence of fever and antibody titers may reflect host intrinsic differences.

There are several limitations to our study that should be noted. First, our cases were all relatively mild (none requiring hospitalization or ventilation), and predominantly middle-aged females. Future studies comparing the responses of older or more severely ill subjects would be informative. Secondly, our study focuses solely on binding of ACE2 to the spike protein and ignores other potential antigens or other immune mechanisms such as inhibition of protease cleavage that might prevent viral entry into the cell. The results of our bead-based assay should be confirmed using a pseudovirus assay, or ideally using live SARS-CoV-2 virus. Third, it is unclear what level of inhibition would correlate with functional resistance to re-infection. Follow-up studies tracking inhibition in our assay over time while simultaneously monitoring subjects for re-infection will be necessary, given the ethical impossibility of experimental human inoculation. Finally, our SARS-CoV-2 PCR+, antibody negative samples leave many questions. For example, did they clear the virus through mechanisms beyond our detection, such as antibodies targeted to alternative viral proteins or non-B-cell dependent mechanisms? Or were their rtPCR results false positives? Most importantly, are they susceptible to future infections? Follow-up with these or similar individuals will be important.

In conclusion, the trimer inhibition assay presented here could be broadly useful in the settings of routine clinical evaluation of functional immunity in recovered patients, selecting the most potent post-convalescent plasma for use as therapy, and evaluating the functionality of antibodies produced in response to experimental vaccines currently under development. In light of our results, we propose that future studies should use trimer-style constructs for serological assays.

## Methods

### Human Samples

Negative controls consisted of banked samples collected from healthy controls prior to January 2020. Samples from negative controls ranged in age from 19 to 66 years of age (median 37) and 18/30 were female (60%). Samples from participants recovered from COVID19 came from the Seattle Children’s SARS2 Recovered Cohort. All subjects reported testing PCR+ for SARS-CoV-2. All parent studies were IRB approved through Fred Hutchison Cancer Research Center or Seattle Children’s Hospital.

### Protein purification

The RBD construct (AA 319-541; UniProt P0DTC2) was cloned into the pCDNA3.4 protein expression vector with an 8XHIS-tag and tandem aviTag. The SARS-CoV-2 trimer^3^ and Ace2 constructs were generously provided by Dr. Jason McLellan (University of Texas at Austin) and Dr. Barney Graham (National Institutes of Health)^3^. Proteins were expressed in 293F cells in antibiotic free, serum free media as previously described^20^. Briefly, DNA was transfected using PEI Max and grown for 5 days at 37C, 5% CO2. The recombinant proteins were purified by NiNTA affinity chromatography and polished by SEC for size on a Superdex 200 16/600.

### CML Bead Coupling

To couple RBD, ACE2 and Trimer proteins to flow cytometry-compatible beads, we followed a previously published protocol^12^. Briefly, 50 μL of carboxylate modified latex (CML) beads (Invitrogen #C37255, USA) were washed 3 times with 500 μL of room temperature MES (50 mM MES (2-(N-morpholino)ethanesulfonic acid), pH 6.0, 1 mM EDTA, in ddH20, stored at 4°C, used at RT) by spinning beads down for 1 minute at 13,000 x g at 4 °C and discarding the supernatant each wash. The CML beads were then resuspended in 50uL of MES and activated with 20 μL of 50mg/mL EDAC (1-ethyl-3-(3-dimethylaminopropyl) carbodiimide HCl; Pierce, USA), freshly dissolved in MES from powder stored at −20 °C. This 70 μL of CML bead, MES, and EDAC was mixed for 15 minutes by gentle, continuous pipetting at room temperature. The beads were washed three times with 0.5 mL of PBS. After the 3rd wash the activated bead pellet was resuspended in a 100 μL solution containing 25 μg of either RBD, Trimer or ACE2 protein in PBS and gently mixed for 3 hours at room temperature at 1400 rpm on a pulsing vortexer. The coupled beads were then washed 3 times in 0.5 mL of PBS and stored in 100 μL of Blocking Storage Solution (1% BSA in PBS, 0.01% Sodium Azide) at 4°C until use. Successful coupling was confirmed by staining ∼0.5 μL coupled beads with PE-conjugated anti-HIS tag antibodies (Santa Cruz, CAT #8036, USA) and compared to positive and negative controls by flow cytometry.

### Magnetic Bead Coupling

MagPlex Microspheres (Luminex #MC100XX-01, USA, where XX encodes the bead region; all regions are compatible) were coupled as previously described^21^. Briefly, 250ul of MagBeads were magnetically separated using a magnetic tube rack (New England Biolabs, Cat# S1506S) for 60 seconds and washed three times with 250ul of MES. MagBeads were resuspended 200ul of MES and 25ul of freshly-prepared 50mg/ml Sulfo-NHS (ThermoScientific, #24510, USA) in MES was added to the beads and briefly vortexed. Then, 25ul of 50mg/ml freshly-made EDAC in MES was added and the tube quickly vortexed. The tube was covered to protect from light and shaken at 1000 RPM for 20 minutes at room temperature on a pulsing vortexer. Beads were then washed twice in PBS, and incubated with of 25ug of Ace2, Trimer or RBD protein in 250ul of PBS, on a 1000 RPM vortexer for 2 hours at room temperature protected from light. Beads were then washed in PBS and quenched in 750 μl of Blocking Storage Solution for 30 minutes at room temperature, 1000 RPM protected from light. Finally, beads were stored in 100 μl of Blocking Storage Solution at 4C until use.

### IgG Detection with IP-FCM

IP-FCM was performed as previously described^11,12^. Patient plasma was obtained from the Seattle Children’s SARS2 Recovered Cohort. Plasma from both SARS-CoV-2 positive and negative patients was heated to 56 °C for 1 hour, then spun at 13,000 x g for 10 minutes at 4 °C. The supernatant plasma was then diluted (1:1000 unless otherwise indicated) in cold Fly-P Buffer (50 mM Tris pH 7.4, 100 mM NaCl, 1% bovine serum albumin, and 0.01% sodium azide) and distributed into wells of a 96-well plate at a volume of 50 μL per well, in duplicate. To each well, approximately 5 x 10^4^ RBD- or Trimer-conjugated CML beads, or 2.5 x 10^3^ MagBeads were added. The wells of the 96-well plate were then capped, and the plate left to rotate at RT for 2 hours. For CML beads, the plate was spun at 3500 RPM to pellet the CML beads, the supernatant was discarded and the beads were washed twice with 125 μl of cold Fly-P Buffer per well. For MagBeads, a magnetic plate washer was used to wash the beads three times (Bio-Plex Pro™ Wash Station, BioRad, USA). The washed beads were incubated with 50 μL of 1:200 anti-Human IgG-PE (Jackson ImmunoResearch, #709-116-149, lot 145536, USA) in Fly-P Buffer protected from light for 30 minutes at room temperature. The plate was washed as above, and samples were resuspended in 50 μL of cold Fly-P Buffer. All plasma samples were also run in parallel using BSA-coupled CML or MagBeads to determine the baseline nonspecific signal generated from each sample. For CML beads, this background varied greatly between individuals from 10^3^-10^5^ MFI units, but was consistent for each individual. MagBeads gave much lower background levels and BSA beads were not used in later experiments. Beads were then read on an Acea Novocyte flow cytometer with the following gating strategy: gate beads using FSC-H vs SSC-H, eliminate doublets using FSC-H vs FSC-A, and detect PE fluorescence using FL2 (488nm excitation, 572/28nm detection). Background-subtracted median fluorescent intensity (MFI) was calculated for each individual by subtracting the BSA-coupled bead MFI from the RBD- or trimer-coupled bead MFI.

### ACE2-Spike binding assay

RBD or trimer protein was biotinylated by adding 1ul of freshly-dissolved sulfo-NHS-Biotin (ThermoScientific, #21217, USA) for 30 minutes on ice. The reaction was quenched with 100mM Tris-HCL and excess biotin was removed by three washes in a 10K MWCO Amicon Ultra spin filter (Millipore, USA). Biotinylated RBD or Trimer protein was added to 5 x 10^4^ ACE2-coupled CML or MagBeads in a total volume of 50 μL, in duplicate. For inhibition assays, soluble unlabeled ACE2 or diluted plasma samples prepared as above were added to each well, maintaining a final reaction volume of 50uL. Plasma was diluted 1:50 in FlyP buffer unless otherwise indicated. Each well of the plate was capped mixed end-over-end at 4 °C overnight. The next day the plate was washed as above and incubated with 50 μL of 1:200 Streptavidin conjugated-PE (BioLegend, #405204, USA) in Fly-P Buffer protected from light for 30 minutes at room temperature. The plate was washed again as above and samples were resuspended in 50 μL of cold Fly-P Buffer for detection on the flow cytometer with gating as above. Data were expressed either as MFI or as percent inhibition, calculated as 1-[MFI of sample]/[MFI of wells without inhibitor added].

### IgG Depletion assay

Patient plasma was diluted 1:50 in Fly-P Buffer and incubated with 2ul of RBD- or trimer-coupled CML beads overnight at 4C. The next day, the beads were pelleted at 13,000 RPM for 1 minute, removed and discarded. A 2ul aliquot of plasma was reserved, and 2ul of new, RBD- or trimer-coupled CML beads were added. This process was repeated for a total of 3 overnight depletions. On the 4^th^ day, 25ul of the RBD- or trimer-depleted plasma was run on the ACE2-trimer binding assay, as described above.

### Statistical Analyses

Technical replicates were averaged and data were imported to Prism 8.0 (GraphPad). Comparisons between two groups were made using two-tailed t-tests, and among three groups with one-way ANOVA followed by post-hoc tests comparing all columns, corrected for multiple comparisons by the Tukey method. Correlations and EC50 values were determined using simple linear regressions or 4-parameter logistic regressions with default settings, and r^2^ values were reported.

## Ethics Statement

All parent protocols were approved by the Fred Hutchinson Cancer Research Center or Seattle Children’s IRBs. This study was conducted under a protocol approved by the Seattle Children’s Research Institute’s IRB STUDY00002514 (SEPS). All participants signed informed consent forms.

## Data Availability

All data are presented in the manuscript.

## Acknowledgements

The authors thank Jason McLellan and Barney Graham for their generous gift of the SARS-CoV-2 recombinant trimer and hACE2 expression constructs; Lee Nelson of the Fred Hutchinson Cancer Research Center for providing negative control samples; Julia Gust for technical assistance; the scientists and support staff at SCRI who enabled COVID research to continue during the pandemic, especially Sean Sandin and Marnie Chinn; and the recovered COVID19+ plasma donors. This study was funded by a COVID-19 research award from the University of Washington Institute for Translational Heath Sciences (via grant UL1 TR002319) and by a COVID19 award from the Research Integration Hub at Seattle Children’s Research Institute (to SEPS), by an NIH/NIAID K08 AI135072 and a Burroughs Wellcome Fund CAMS 1017213 grant (to WH), and by R01 AI140951 (to DNS).

## Conflict of Interest

The authors declare no conflicts of interest. A provisional patent application has been filed by SCRI on the assays reported in this paper.

## Author Contributions

SEPS conceived the study. EPG, SEPS, NS and WH planned experiments. EPG performed all experiments except those noted as follows. ND, VV, and OT designed, expressed and purified recombinant proteins and performed QC and antigenic properties. YJ, WH and LF conducted the Seattle Children’s SARS2 Recovered Cohort sample collection and provided samples and deidentified patient data. EPG, SEPS, WH and DNS analyzed the data. EPG, SEPS, WH and LF wrote the manuscript, and all authors read and approved the manuscript.

---

**Figure S1:**
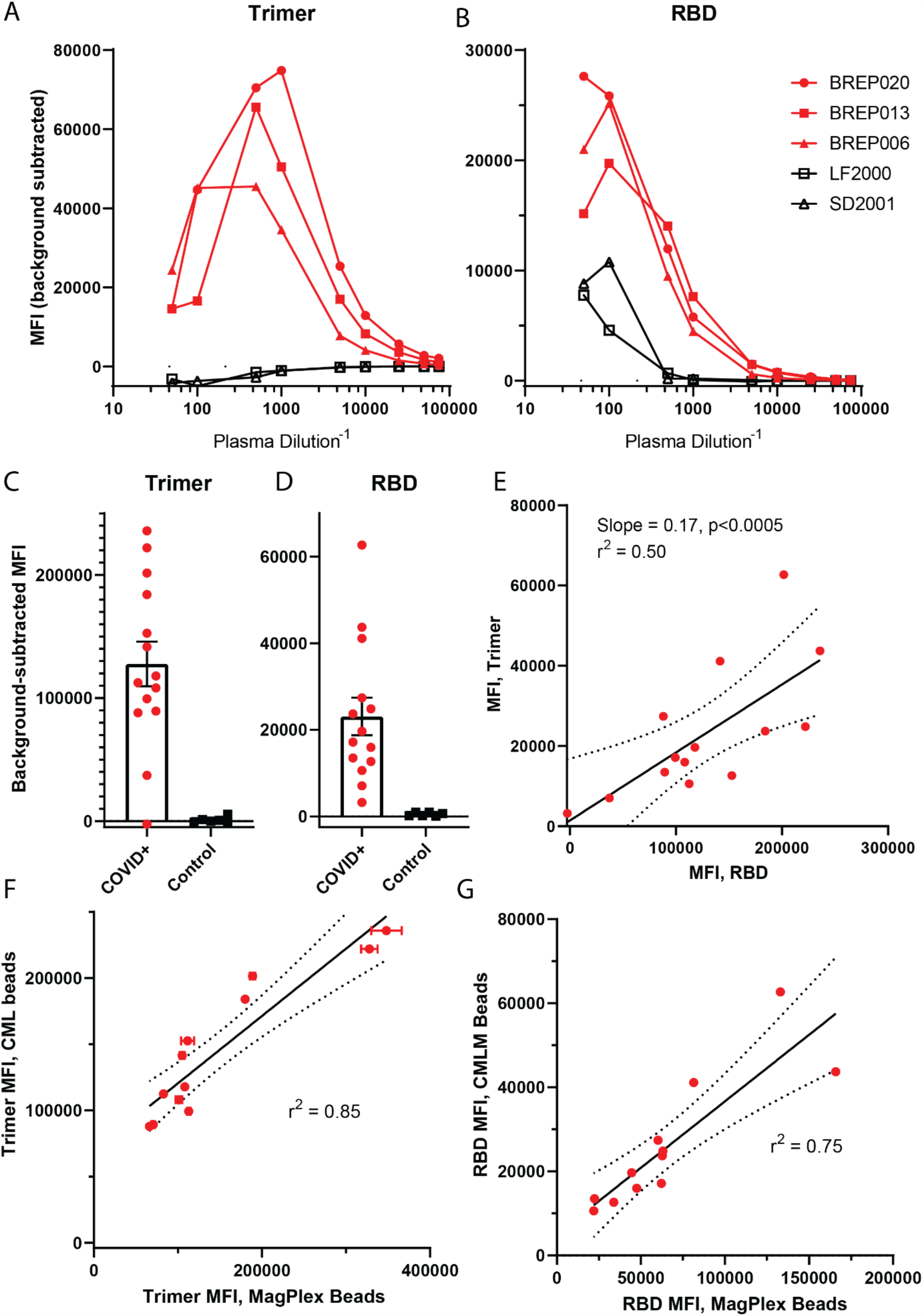
Related to Figure 1. Detection of anti-SARS-CoV-2 IgG using RBD and trimer constructs using CML beads. (A-B) Plasma from 3 COVID+ samples (red) and two controls (black) was serially diluted, and IgG levels were measured using trimer-conjugated (A) or RBD-conjugated (B) microspheres. (C-D) Plasma from 14 COVID+ and 6 pre-covid controls was diluted 1:1000, and IgG levels were measured using trimer-conjugated (C) or RBD-conjugated (D) microspheres. E) The median fluorescent intensity of IgG measured on the trimer vs. RBD assays was significantly correlated. Solid line shows a linear regression with 95% confidence intervals indicated with dotted lines, r^2^ = 0.50. (F) Anti-trimer IgG measured by the CML bead assay (as in Figure S1C) plotted vs. anti-trimer IgG measured by the MagBead bead assay (as in Figure 1B) for the 13 COVID+ samples measured on both assays. Solid line shows a linear regression with 95% confidence intervals indicated with dotted lines, r^2^ = 0.85. (G) Anti-RBD IgG measured by the CML bead assay (as in Figure S1D) plotted vs. anti-RBD IgG measured by the MagBead bead assay (as in Figure 1C) for the 13 COVID+ samples measured on both assays. Solid line shows a linear regression with 95% confidence intervals indicated with dotted lines, r^2^= 0.75.

**Figure S2:**
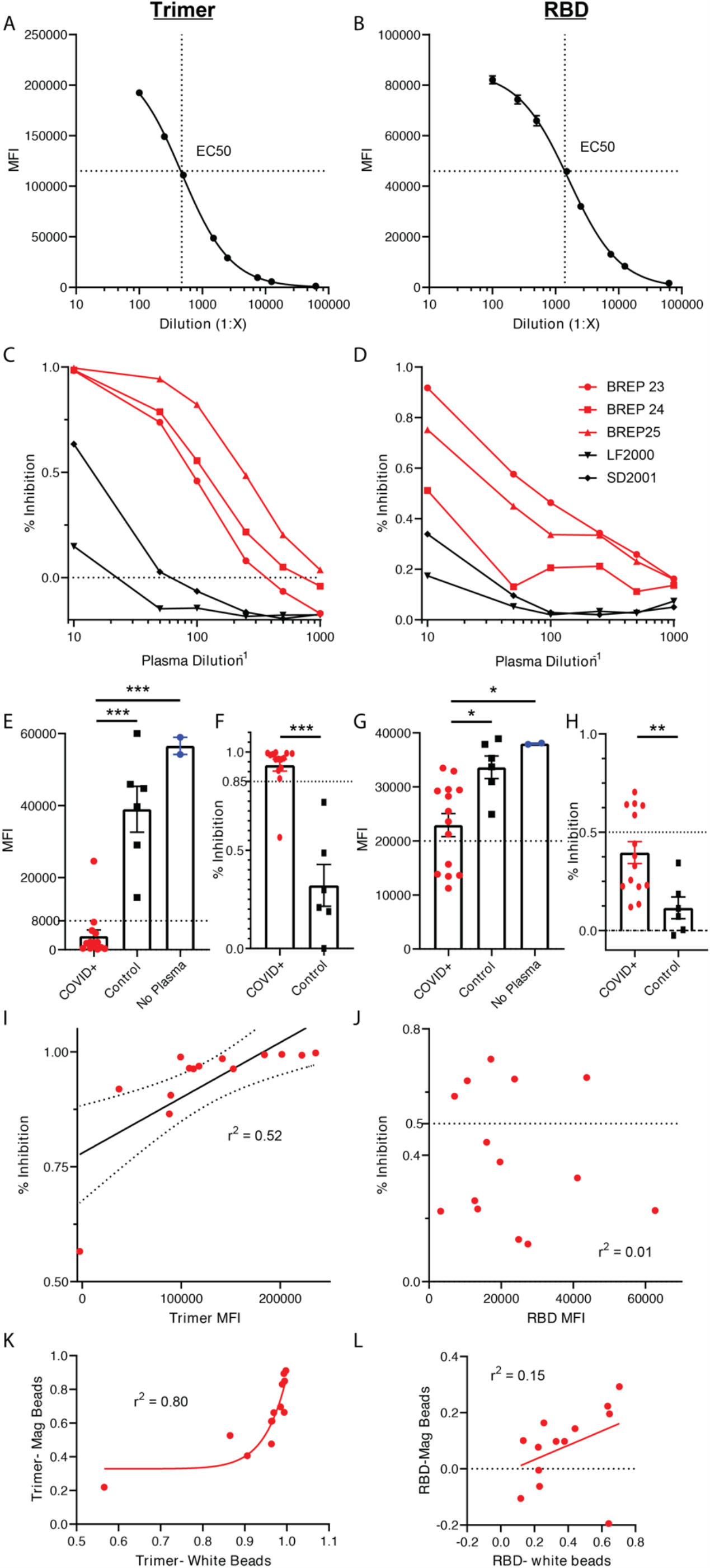
Related to Figure 2. Inhibition of SARS-CoV-2 spike protein and Ace2 binding by recovered patient plasma using CML beads. (A,B) The EC50 of the Ace2-Trimer (A) or Ace2-RBD (B) binding reaction was determined by serial dilution of the trimer or RBD fragment. (C,D) Plasma from 3 COVID+ samples (red) and two controls (black) was serially diluted, and inhibition of Ace2 binding was measured using the trimer (C) or RBD (D) inhibition assay. E) Compared to a no-plasma control (blue), pre-COVID control plasma significantly inhibited Trimer-ACE2 binding (p<0.05), but COVID+ plasma inhibited binding to a much greater degree (P<0.001 vs control plasma and no plasma). F) Same data as in A expressed as % inhibition vs. the no-plasma control average. An arbitrary cut-off of 85% inhibition captures all IgG-positive COVID samples. P<0.001. G) Compared to a no-plasma control (blue), pre-COVID control plasma slightly and non-significantly inhibited RBD-ACE2 binding. COVID+ plasma inhibited in some samples, such that the group mean was significantly different from both pre-COVID and no plasma controls (P<0.005 vs control plasma and p<0.01 vs no plasma). However, the population appeared to bifurcate into a responder and non-responder population (dotted line at MFI = 20,000). H) Same data as in C expressed as % inhibition vs the no-plasma control average. P<0.005. I) Trimer MFI plotted vs. % inhibition on the Trimer-Ace2 inhibition assay for all COVID+ samples. J) RBD MFI plotted vs. % inhibition on the RBD-Ace2 inhibition assay for all COVID+ samples. K) % inhibition on the trimer-Ace2 inhibition assay on MagBeads plotted vs. the same assay run on CML beads, for the 12 individuals that were run on both assay types. L) % inhibition on the RBD-Ace2 inhibition assay on MagBeads plotted vs. the same assay run on CML beads, for the 12 individuals that were run on both assay types.

## Notes

### Competing Interest Statement

The authors have declared no competing interest.

### Author Declarations

All parent protocols were approved by the Fred Hutchinson Cancer Research Center or Seattle Childrens IRBs. This study was conducted under a protocol approved by the Seattle Childrens IRB STUDY00002514 (SEPS).

## References

1. Huang, C. et al.. Clinical features of patients infected with 2019 novel coronavirus in Wuhan, China. The Lancet 395, 497–506 (2020).

2. Cucinotta, D. & Vanelli, M. WHO Declares COVID-19 a Pandemic. Acta Bio-Medica Atenei Parm. 91, 157–160 (2020).

3. Wrapp, D. et al.. Cryo-EM structure of the 2019-nCoV spike in the prefusion conformation. Science 367, 1260–1263 (2020).

4. Walls, A. C. et al.. Structure, Function, and Antigenicity of the SARS-CoV-2 Spike Glycoprotein. Cell 181, 281-292.e6 (2020).

5. Tai, W. et al.. Characterization of the receptor-binding domain (RBD) of 2019 novel coronavirus: implication for development of RBD protein as a viral attachment inhibitor and vaccine. Cell. Mol. Immunol. 1–8 (2020) doi:10.1038/s41423-020-0400-4.

6. Letko, M., Marzi, A. & Munster, V. Functional assessment of cell entry and receptor usage for SARS-CoV-2 and other lineage B betacoronaviruses. Nat. Microbiol. 5, 562–569 (2020).

7. Chen, X. et al.. Human monoclonal antibodies block the binding of SARS-CoV-2 spike protein to angiotensin converting enzyme 2 receptor. Cell. Mol. Immunol. 1–3 (2020) doi:10.1038/s41423-020-0426-7.

8. Long, Q.-X. et al.. Antibody responses to SARS-CoV-2 in patients with COVID-19. Nat. Med. 1–4 (2020) doi:10.1038/s41591-020-0897-1.

9. Wu, F. et al.. Neutralizing antibody responses to SARS-CoV-2 in a COVID-19 recovered patient cohort and their implications. medRxiv 2020.03.30.20047365 (2020) doi:10.1101/2020.03.30.20047365.

10. Wang, C. et al.. A human monoclonal antibody blocking SARS-CoV-2 infection. Nat. Commun. 11, 2251 (2020).

11. Smith, S. E. P. et al. IP-FCM Measures Physiologic Protein-Protein Interactions Modulated by Signal Transduction and Small-Molecule Drug Inhibition. PLOS ONE 7, e45722 (2012).

12. Davis, T. R. & Schrum, A. G. IP-FCM: Immunoprecipitation Detected by Flow Cytometry. JoVE J. Vis. Exp. e2066 (2010) doi:10.3791/2066.

13. Amanat, F. et al.. A serological assay to detect SARS-CoV-2 seroconversion in humans. Nat. Med. (2020) doi:10.1038/s41591-020-0913-5.

14. Kirkcaldy, R. D., King, B. A. & Brooks, J. T. COVID-19 and Postinfection Immunity: Limited Evidence, Many Remaining Questions. JAMA (2020) doi:10.1001/jama.2020.7869.

15. Xing, Y. et al.. Post-discharge surveillance and positive virus detection in two medical staff recovered from coronavirus disease 2019 (COVID-19), China, January to February 2020. Eurosurveillance 25, 2000191 (2020).

16. Bao, L. et al.. Lack of Reinfection in Rhesus Macaques Infected with SARS-CoV-2. bioRxiv 2020.03.13.990226 (2020) doi:10.1101/2020.03.13.990226.

17. KCDC. KCDC. KCDC http://www.cdc.go.kr.

18. To, K. K.-W. et al. Temporal profiles of viral load in posterior oropharyngeal saliva samples and serum antibody responses during infection by SARS-CoV-2: an observational cohort study. Lancet Infect. Dis. 20, 565–574 (2020).

19. Zhao, J. et al. Antibody responses to SARS-CoV-2 in patients of novel coronavirus disease 2019. Clin. Infect. Dis. doi:10.1093/cid/ciaa344.

20. Fisher, B. S. et al. Oral Immunization with HIV-1 Envelope SOSIP trimers elicits systemic immune responses and cross-reactive anti-V1V2 antibodies in non-human primates. PloS One 15, e0233577 (2020).

21. Brown, E. A., Neier, S. C., Neuhauser, C., Schrum, A. G. & Smith, S. E. P. Quantification of Protein Interaction Network Dynamics using Multiplexed Co-Immunoprecipitation. JoVE J. Vis. Exp. e60029 (2019) doi:10.3791/60029.

